# Protocol for a Safety, Feasibility, and Dose-Finding Study of the Hemopurifier® Device in Patients with Solid Tumors Who Have Stable or Progressive Disease During Pembrolizumab or Nivolumab Monotherapy

**DOI:** 10.1101/2025.03.20.25323761

**Authors:** Michael P. Brown, Marco Matos, Stephen Clarke, P. Toby Coates, Carol Pollock, Jagadeesh Kurtkoti, Georges Grau, Kelly Edinburg, Alessio Bloesch, Rosalia de Necochea Campion, Nyan Y. Khin, John Shin, Steven P. LaRosa

**Author notes:** **Corresponding Author:** Steven P. LaRosa, M.D. **Funding sources** Funding for this study was provided by Aethlon Medical Australia PTY LTD.

## Abstract

**Background:** Immunotherapy with anti-PD-1 inhibitors has revolutionized the treatment of many solid tumors, however only 30-40% of patients will have a lasting clinical response. Tumor-derived extracellular vesicles (EVs) have been implicated in the spread of solid tumors and resistance to these agents. A lectin-affinity plasmapheresis device called the Hemopurifier (HP) has been developed and shown to remove extracellular vesicle in vitro and in patients. We hypothesize that the treatment of patients who are not improving on anti-PD-1 therapies will be safe, decrease EV concentrations, and improve anti-tumor T cell activity.

**Methods:** This safety, feasibility and dose finding study is designed in a 3+3 safety study design with three treatment cohorts. Participants who are determined to be not responding to anti-PD-1 monotherapy will be assigned to receive either one, two, or three (HP) treatments over a one-week period prior to their next scheduled dose of anti-PD-1 antibody. Advancement from one cohort to the next will be determined by a DSMB. Data collection will include adverse events, safety labs, EV concentrations and T cell measurements, repeat imaging and survival status.

The primary outcome of the study will be the safety of the HP in this population with additional endpoints to include the kinetics of EV removal and rebound following HP treatment, in addition to the effects on T cell numbers and activity,

**Ethics and dissemination:** The clinical protocol and amendment to the study protocol have been approved by the Ethics Committees and Research Governance Offices at three participating clinical sites in Australia. The Therapeutic Good Authority (TGA) has been notified. The clinical trial is listed on the Australia New Zealand Clinical Trials Registry. Informed Consent is obtained from all participants prior to any protocol procedures being performed.

Australia New Zealand Registration number ACTRN12624000732583

## INTRODUCTION

Immunotherapy, which attempts to harness the immune system to attack malignant cells has been a game-changer in the treatment of many solid tumors. One target for inhibition has been immune checkpoints. It is estimated that approximately 46% of cancer patients in the United States are eligible for these agents ^1^. One such checkpoint is Program Death Ligand −1 (PD-L1). PD-L1 expressed by cancer cells binds to the PD-1 receptor on lymphocytes rendering them inactive. Anti-PD-1 monoclonal antibodies including pembrolizumab and nivolumab have been designed to prevent T cell inactivation by PD-L1 ^2^. Pembrolizumab, an anti-PD-1 inhibitor, has garnered 40 FDA approvals ^3^and is estimated to have 2 million commercial patients by the end of 2024.

Despite the success of anti-PD-1 therapies, longstanding clinical response is seen on only approximately 30-40% of patients ^4^. Extracellular vesicles (EVs) have been implicated in the spread of cancers as well as resistance to anti-PD-1 agents ^5,6^. EVs including small 50-200 nm particles called exosomes are nanoparticles with a lipid bilayer released by all cell types including cancer cells. EVs are involved in cell-to-cell communication modulating the local and distant microenvironment through cargo including metabolites, proteins, microRNAs, DNAs and RNAs. This cargo promotes tumor growth and invasion, metastasis, and angiogenesis. Extracellular vesicles also participate in chemotherapy resistance through mechanisms including drug expulsion ^5^.

Immune suppression mediated by EVs plays a role in cancer pathogenesis. Additionally, EVs can carry PD-L1 on their surface allowing them to bind to PD-1 on lymphocytes and inactivate them ^6^. Sustained levels of exosomal PD-L1 during therapy with anti-PD-1 agents have been associated with poor response to anti-PD-1 therapies ^7,8^. This raises the hypothesis that removing EVs could restore responsiveness to anti-PD-L1 antibodies. Efforts have been made to block exosomal biogenesis with small molecules ^9^A complete blockade of EVs that carry beneficial cargo could however result in an adverse safety profile ^10^. A strategy of “debulking” rapidly produced tumor-derived EVs rather than a complete blockade of all EV production may allay this concern.

A lectin-affinity plasmapheresis device called the Hemopurifier® (HP) (Aethlon Medial, Inc., San Diego, CA, USA) has been developed to target and remove EVs in Oncology. The device is a single-use plasma separator cartridge that is modified to contain a proprietary affinity resin (consisting of the lectin *Galanthus nivalis* agglutinin (GNA) attached covalently to the silica in diatomaceous earth) in the plasma space, external to hollow fibers with a nominal pore size of 200-500nm ^11^. The lectin GNA has been found to bind to terminal non-reducing or reducing mannose as well as alpha-1,3 linked mannose oligosaccharides ^12^. Glycan analysis in exosomes derived from melanoma cell lines and ovarian cell lines has revealed that these tumors selectively express high mannose type glycans on their surface ^13,14^.

Blood is accessed from the patient utilizing a standard dual lumen vascular catheter and a dialysis machine as a blood pump. Blood enters the cartridge where plasma is forced through the pores of the hollow fiber membrane because of the pressure gradient established. Blood cellular elements remain in the lumen of the hollow fibers while plasma enters the space outside the fibers where the lectin-affinity resin resides. Mannosylated molecules are bound by the lectin and prevented from re-entering the circulation. As the pressure gradient dissipates along the course of the hollow fibers plasma re-enters the lumen and joins the cellular elements to be returned to the patient without the need for plasma replacement^11^.

Both *in-vitro* and *in-vivo* binding of EVs by the Aethlon HP has been demonstrated. Two in vitro studies have been conducted utilizing a benchtop version of the device demonstrating EV removal in plasma samples from cancer patients. One of these studies examined EV removal from buffer and the second examined EV removal directly from plasma.

In the first *in vitro* experiment utilizing a circuit and a benchtop version of the HP, EVs were isolated from plasma of cancer patients from tumor types including head and neck cancer, melanoma, ovarian cancer, esophageal cancer, and breast cancer. These EVs were suspended in buffer and recirculated over the benchtop device packed with the affinity resin. EVs remaining in the buffer were quantified using nanoparticle tracking analysis measurements and the ExoView instrument. The results demonstrated 92-99% clearing of EVs from input concentrations of 10^9^to 10^10^EVs per mL ^15^.

In a recently completed second *in vitro* experiment, 44 mL of frozen plasma from a patient with non-small cell lung cancer, who had received treatment with a variety of therapies including the anti-PD-1 antibody nivolumab, was diluted with non-cancer patient plasma to a final volume of 50mL. This volume was circulated via a pump over a pediatric HP device (1/12^th^the capacity of a full-sized HP) loaded with the affinity resin for the number of passes that would equate to a 4-, 6- and 24-hour treatment for a normal adult plasma volume over an adult size HP. Plasma samples were taken at baseline and at three time points (follow-ups 1, 2 and 3, respectively) for EV quantification. Quantification of EVs was performed by Cellarcus Labs (San Diego, CA, USA) utilizing vesicle flow cytometry employing a lipid membrane dye (vFRed) and either labeled-Annexin V or tetraspanin antibodies to CD9/CD63, and CD81 as EV-selective stains.

Determinations were performed directly on the plasma samples and separately on Size Exclusion Chromatography (SEC) (isolates of EVs from the plasma from which most non-vesicular nanoparticles (NVLPs) have been removed. At follow-up #2 (equating to 6 hours of HP treatment) exosome concentrations determined by Tetraspanin had decreased by 31% in the raw plasma samples and 46% in SEC prepared samples. At follow-up #2 (equating to 6 hours of HP treatment) exosome concentrations determined by Annexin V had decreased by 66 % in the raw plasma samples and 62% in SEC isolates.

*In vivo* reduction of EVs was measured during a total of ten HP sessions in 2 patients. Eight of the ten treatment sessions occurred in a single patient with severe COVID-19 infection and multi-organ system failure without malignancy^11^. A downward trajectory in total exosome concentration was observed over the course of the 8 treatments. Additionally, when exosomal cargo was examined, it was found that HP treatment was associated with decreases in exosomal miRNA-424, which is associated with COVID-associated coagulopathy and exosomal miRNA-16 which is associated with acute lung injury.

A previous US study in head and neck cancer patients conducted during the pandemic yielded data on EV removal by the HP (previously unpublished data). A single patient with head and neck cancer underwent two 4-hour HP sessions separated by 21 days. EV concentrations were measured via Nanosight before the treatment and on days 1,2,3,4,7 and 14 following each treatment. The total nanoparticle counts decreased following each of the treatments and slowly began to rise 7 days after each treatment to levels that were below the baseline measurement (Figure 1).

**Figure 1.**
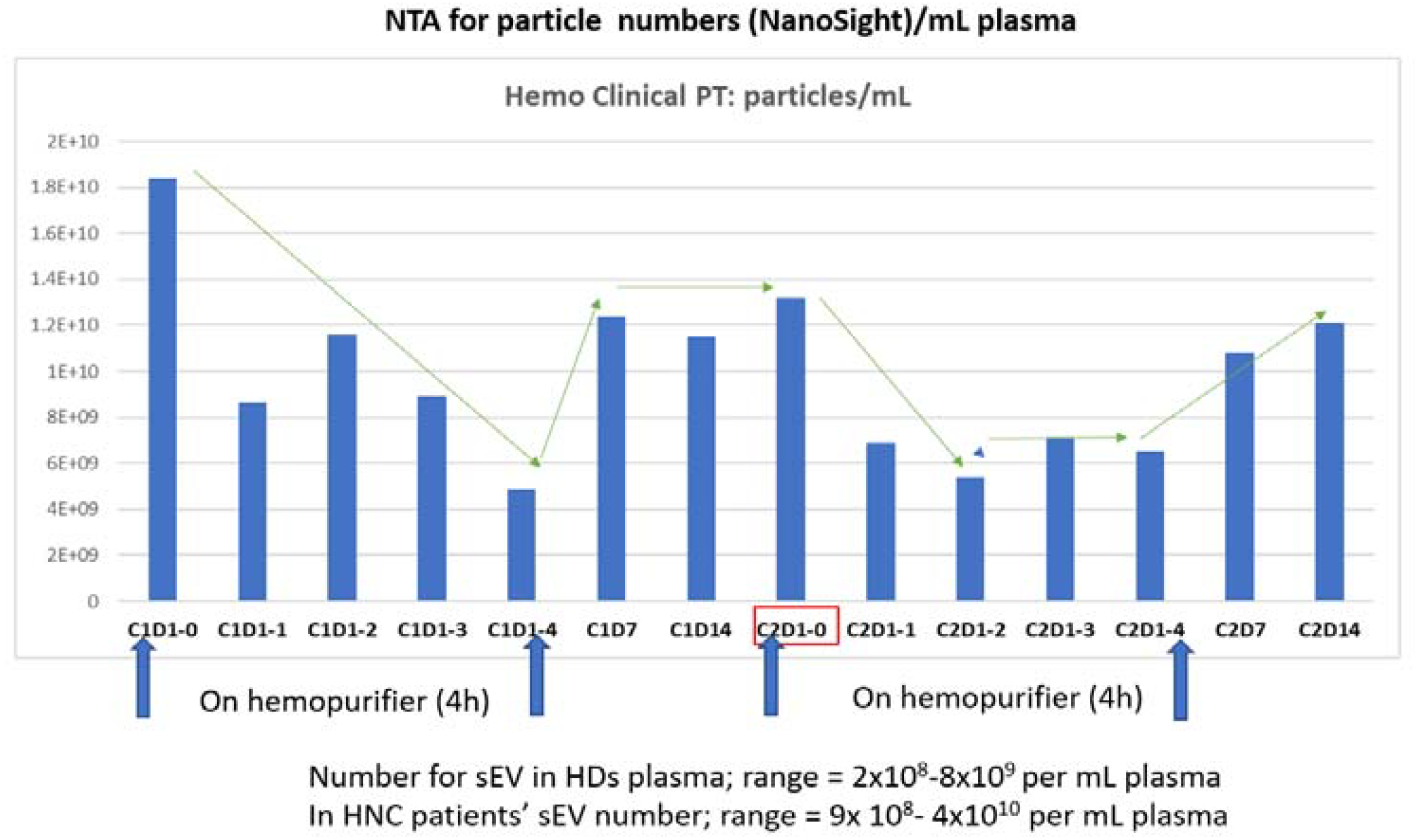
Nanoparticles counts following HP treatment in a patient with head and neck cancer.

The safety profile of the HP has been examined to date in 165 distinct HP sessions completed by a total of 39 patients. Patients treated included those with HCV infection, HIV infection, Ebola infection, COVID-19 infection and two patients with malignancy. These HP treatments have been well-tolerated. In early studies hemolysis was observed when blood flow rates were more than 200 ml/min (current blood flow rates are maintained at 150ml/min). Additional adverse events (AEs) have included shivering, fever, nausea, dry cough, backache, sore throat, itching, and blood leaks. One patient who had received an angiotensin-converting enzyme (ACE) inhibitor (an excluded criteria in the clinical trial) experienced an anaphylactoid reaction.

The current study seeks to examine the safety and feasibility of the HP in several tumor types in patients who are not responding to anti-PD-1 therapy. An important additional aim of the study is to determine the frequency of HP sessions necessary to lead to sustained decreases in EV concentrations. A pilot clinical study of total plasma exchange in patients who required plasmapheresis indicated that three treatments in a week were necessary to achieve sustained decreases in exosomal PD-L1 levels ^16^. Tumor markers will be employed to study the relative removal of tumor-derived vs non-tumor derived EVs in different tumor types by the Hemopurifier. We will also explore the effects of decreases in EV concentrations on anti-tumor T cell activity during the trial. An improvement in the T cell proliferation as indicated by a rise in Ki-67^+^PD-1^+^CD8 cells, has been proposed as a marker of response to anti-PD-1 antibody therapy ^17^. If successful, the data from this trial will inform the design of a subsequent larger safety and efficacy trial.

## METHODS

### Overview of Study Design

This feasibility study will assess:

1. the safety of the HP in patients with solid malignancies not responding to anti-PD-1 therapies (primary outcome),
2. the kinetics of EV removal and rebound following HP treatment as well as the effects on T cell activity and
3. the radiographic response by RECIST 1.1 criteria at 10-12 weeks post HP treatment and one-year survival.

### Recruitment, screening and treatment arms

Patients with solid tumors who are deemed by their treating physician to have stable or progressive disease while on monotherapy with either pembrolizumab or nivolumab are eligible for this study. Full inclusion and exclusion criteria are found in Table 1. Potentially eligible patients will be approached by clinical trial coordinators and will provide written informed consent prior to the performance of any screening procedures.

**Table 1.** Inclusion and Exclusion criteria.

| Inclusion Criteria | Exclusion Criteria |
| --- | --- |
| 1. Histologic diagnosis of one of the following solid tumours:<br>• NSCLC<br>• Melanoma<br>• Urothelial<br>• Renal<br>• Colorectal cancer (MSI-H or dMMR)<br>• Gastric, GEJ, or oesophageal<br>• Head and neck<br>• Cervical<br>• Mesothelioma<br>2. Stable or progressive disease during treatment with pembrolizumab or nivolumab monotherapy<br>3. ECOG PS: 0-1<br>4. Measurable disease by RECIST 1.1<br>5. ≥18 years of age<br>6. Ability to provide informed consent<br>7. Life expectancy of at least 12 weeks<br>8. Adequate contraception (WOCBP/men, up to 120 days post HP treatment) | 1. Presence of brain or leptomeningeal metastasis<br>2. Immunodeficiency disorder<br>3. Any autoimmune condition requiring immunosuppressive treatment including oral or intravenous corticosteroids within 12 months prior to enrolment<br>4. Currently being treated for acute non-infectious pneumonitis<br>5. Pregnant or breastfeeding (WOCBP must have a negative urine pregnancy test performed on the same day but prior to the first HP treatment).<br>6. HIV infection<br>7. Active hepatitis B or C infection<br>8. Use of an angiotensin-converting enzyme (ACE) inhibitor within 14 days prior to a HP treatment.<br>9. Systolic blood pressure <100 mmHg during Screening Period in a patient with a history of a systolic blood pressure ≥ 100 mmHg<br>10. Contraindication to anticoagulation: unable to tolerate full therapeutic anticoagulation therapy for any reason<br>11. Hemoglobin < 90g/L<br>12. Absolute Neutrophil count < $1.5 \times 10^9$ /L<br>13. Platelet count < $75 \times 10^9$ /L<br>14. Creatinine Clearance < 45 ml/min (2021 CKD-EPI)<br>15. Serum albumin ≤ 30g/L<br>16. Bilirubin ≥ 25.66 µmol/L<br>17. Aminotransferases levels more than five times above the Upper Limit of Normal (×5 times the ULN)<br>18. Any disorder where the patient would not tolerate placement of a venous-venous vascular catheter, blood volume loss during an extracorporeal session or from research blood draws per the judgment of the principal investigator<br>19. Patient refuses to receive blood replacement if required<br>20. History of allergy to heparin or heparin-induced thrombocytopenia<br>21. History of solid organ transplantation<br>22. Any condition which, in the opinion of the investigator, makes the patient a poor candidate for this clinical trial |
|  | 23. Patients may not participate in any other investigational trial while participating in this study |

Participants who successfully complete screening will sequentially be assigned to cohorts of 3 participants to receive either 1, 2, or 3 HP treatments over a one-week period prior to their next dose of pembrolizumab or nivolumab (Figure 2).

**Figure 2.**
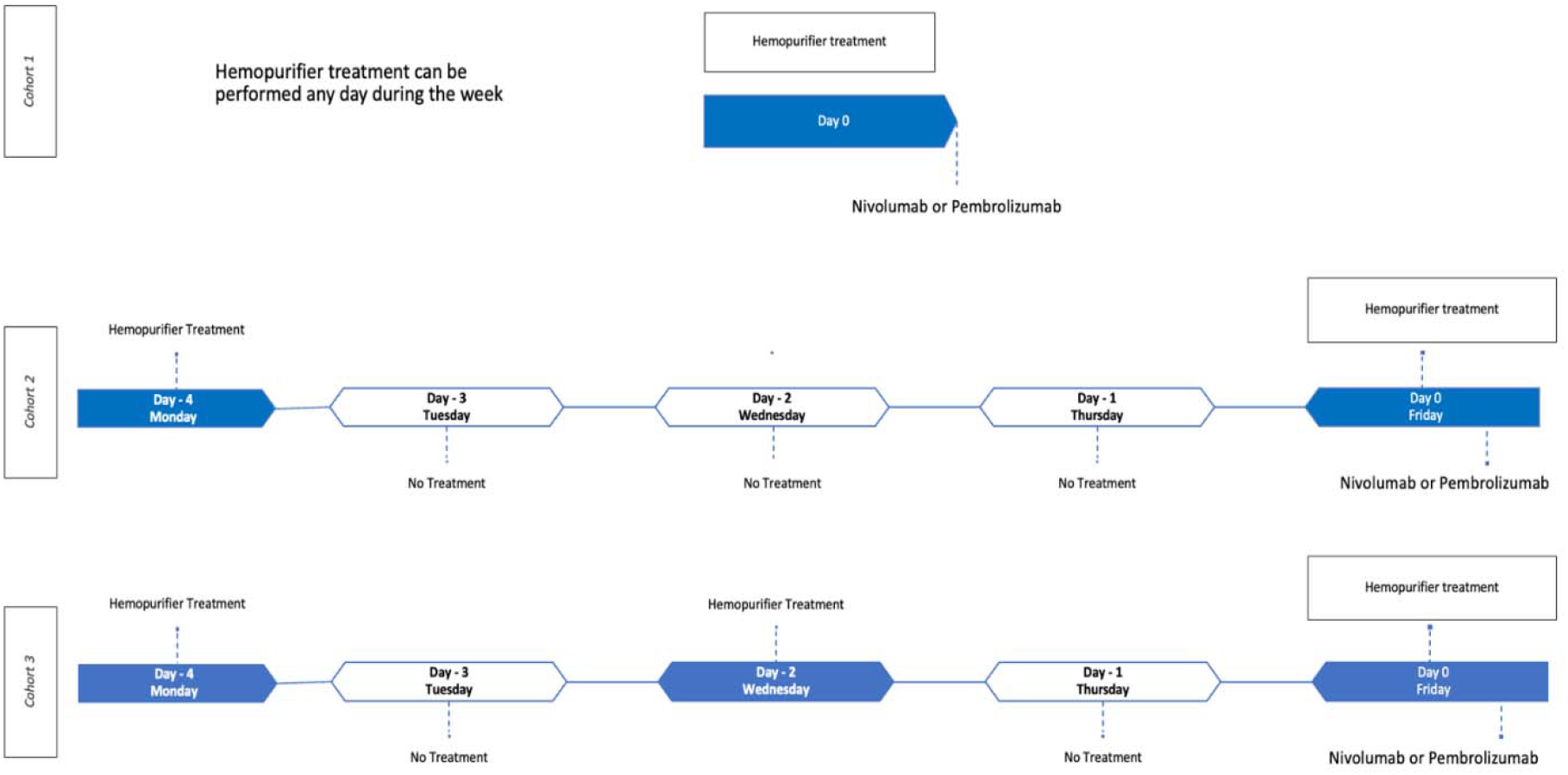
Treatment Cohorts. The day(s) of scheduled HP treatment in each cohort are highlighted blue.

The study will employ a 3+3 safety design whereby 3 participants will be added to a cohort if a participant develops a dose limiting toxicity (DLT) or serious adverse event that is considered unexpected and device related. If two participants in a cohort develop one of these criteria the trial will be terminated. If a participant does not complete the prespecified 7-day safety follow-up after HP treatment they will be withdrawn from the study and replaced by another participant in the same cohort. Based on this design it is anticipated that anywhere from 2 to a maximum of 18 participants may be enrolled. An independent Data Safety Monitoring Board (DSMB) will review the 7-day safety follow-up data from each cohort to determine advancing from one cohort to the next.

### Study Interventions

All participants will continue to receive their anti-PD-1 therapy throughout the study, with the dose, dosing interval and duration at the discretion of the treating physician. All participants will receive their usual concomitant medications for any underlying medical conditions without interruption. Participants may receive radiotherapy but not any other concomitant anti-cancer agent from enrollment to 8 weeks after the last HP treatment. Participants may not receive ACE inhibitors within 14 days prior to, or during their HP treatment(s). Participants will not receive HP treatment if they have been treated with corticosteroids for autoimmune conditions or immune-related adverse events in the 12 months prior to HP treatment.

The participant will have a double lumen veno-venous catheter placed under radiologic guidance. The HP treatment will be conducted for 4 hours at a target flow rate of 150-170 ml/min delivered by an approved dialysis machine. Vital signs, cardiac monitoring, pulse oximetry, and venous pressure and arterial pressure on the dialysis machine will be monitored and recorded throughout the treatment. In addition, a post-pump pre-HP pressure monitor will be used as an early monitoring system to detect clotting within the device. Visual inspection of the device will be performed to assess for clotting, hemolysis, and broken fiber blood leaks throughout the treatment. The participant will receive either unfractionated heparin or low molecular weight heparin throughout the HP treatment to achieve therapeutic anticoagulation. Point of care Activated Clotting Time (ACT) measurements will be used to ensure adequacy of anticoagulation. The vascular catheter will be removed following the last HP treatment following decreases in ACT measurements to safe levels.

### Data Collection Schedule

Prior to the 1^st^HP treatment, the participant will have samples taken for baseline measurements of hematology, chemistry, EV concentration and T cell numbers and activity. Imaging will be performed for measurement of disease burden by RECIST 1.1.

Samples will be taken 2 hours into the HP treatment and at the end of treatment at each session for determination of EV concentrations. In cohorts 2 and 3 samples will be taken on the days in between HP treatments for the determination of EV concentrations.

All participants will return to the clinic on weeks 1, 2, 3, 4 and 8 following the last HP treatment. Items to be performed at these visits will include determination of adverse events, physical exam, ECOG performance status, safety labs, and lab samples for determinations of EV concentrations and T cell activity. At 10-12 weeks post the last HP treatment imaging will be repeated for follow-up RECIST 1.1 scoring. Participants will have follow-up at 26 and 52 weeks for adverse event collection and determination of survival status.

### Measuring EV Numbers and Phenotypes

#### Processing of plasma samples

Plasma will be processed and frozen in aliquots at −80°C to be shipped to the central laboratory. The frozen aliquots, consisting of platelet-poor plasma, are thawed and centrifuged at 18,000 g for 3 minutes. The final supernatant collected will be referred to as platelet-free plasma (PFP). All steps are conducted at room temperature.

#### EV enrichment

200 µL PFP will be collected and diluted with 800 µL of 0.02 µm filtered PBS before undergoing centrifugation at 18,000 g for 45 minutes at 4°C. This “18 k pellet” will be resuspended with 350 µL of filtered PBS to collect microvesicles (MV), now called large EV (lEV). To collect exosomes (now called small EV, sEV), 950 µL of the 18 k supernatant will be collected and further diluted with 500 µL of 0.02 µm filtered PBS before undergoing centrifugation at 150,000 g for 2 hours at 4°C. The pellet obtained this way will be called “150 k pellet”.

#### Flow cytometry

20 µL of the MVs/lEV preparation will be stained with fluorescently conjugated antibodies (anti-CD3-AF532 (Invitrogen), anti-CD41-APC (BioLegend), anti-CD9-PE (Invitrogen), anti-CD63-AF488 (BioLegend), anti-EGFR-FITC (BioLegend), anti-MUC1-PE (BioLegend), anti-PD1-AF488 (BioLegend)), or Annexin V-FITC (Beckman Coulter) for 20 minutes in the dark. After incubation, 180 µL of 0.02 µm filtered PBS will be added before acquisition on an Aurora 3L flow cytometer (Cytek Biosciences). Samples with higher than 8,000 events will be further diluted with PBS to ensure the number of vesicles can be accurately determined. Results will be analyzed using FlowJo software version 10.

#### Nanoparticle tracking analysis

1400 µL supernatant of the 150 k pellet will be removed and the 150 k pellet will be resuspended with 150 µL of filtered PBS to collect exosomes/sEV. The exosomes will be further diluted 1:500 with 0.02 µm filtered PBS. Fluorescently conjugated antibodies (anti-CD3-AF532 (Invitrogen), anti-CD41-APC (BioLegend), anti-CD9-PE (Invitrogen), anti-MUC1-PE (BioLegend), anti-EGFR-AF647 (BioLegend), anti-EPCAM-AF647 (BioLegend), anti-CD147-AF647 (BioLegend)) will be added to 9 µL of this dilution for 30 minutes in the dark, followed by another addition of 990 µL of filtered PBS. The sample will then be injected via a syringe into the ZetaView QUATT (Particle Metrix) to phenotype and determine the numbers and sizes of exosomes/sEV.

### Measuring T cell numbers and Activity

#### PBMC isolation

PBMC will be isolated using the EasySep Direct Human PBMC Isolation Kit (Stemcell; 19654) according to the manufacturer’s instructions. Briefly, whole blood collected in Cyto-Chex BCT tube is transferred to a 14 mL round bottom tube and incubated with EasySep Isolation Cocktail (1:20 dilution) for 5 mins. The sample is diluted 1:1 with EasySep Buffer (Stemcell; 20144) followed by the addition of EasySep Rapid Spheres at 1:20 dilution. This mixture is placed into the Big EasySep Magnet (Stemcell; 18001) for 5 minutes. This cell suspension is then poured into a new tube and incubated again with EasySep Rapid Spheres for 5 minutes on the magnet. The cell suspension is poured out and immediately placed on the magnet for 5 minutes with no Rapid Spheres. The final cell suspension is poured into a 15 mL Falcon tube and viable PBMC counts are recorded. Cells will be analyzed by flow cytometry or stored in liquid nitrogen for later analyses. All steps will be performed at room temperature.

#### Flow cytometry

Human PBMC will be blocked with Anti-human Fc Receptor Binding Inhibitor (Invitrogen; 14-9161-73) in FACS buffer prepared with DPBS, 0.5% Bovine Serum Albumin (BSA) and 2 mM EDTA for 10 minutes. Cells are then stained with primary conjugated antibodies (Table 1) diluted in FACS buffer for 20 minutes. After staining, cells will be fixed with 4% paraformaldehyde for 20 minutes and assessed using an Aurora 5L spectral flow cytometer. All steps will be performed at room temperature and results analyzed using FlowJo software version 10.

### Study Objectives and Endpoints

The primary objective of the study is to assess the safety and feasibility of the HP dosed at different intervals in patients with solid tumors with stable or progressive disease as determined by the treating physician during pembrolizumab or nivolumab monotherapy.

The secondary objectives of the study include assessing the concentration of total EVs as well as cancer and host derived EVs and EV cargo during HP therapy, to assess the concentration of EV cargo and EV subsets during HP therapy and to assess the rebound of EV and EV cargo over time at different HP dosing intervals. An exploratory objective is to assess the effects of HP therapy dosed at different intervals on T cell numbers and activity.

### Statistical Methods

A full Statistical Analysis Plan (SAP) will be prepared to expand on the information provided below. The sample size for the study is based on safety and not based on any statistical hypothesis. A 3+3 dose escalation design has been chosen. Based on this design it is anticipated that anywhere from 2 to a maximum of 18 participants will be enrolled. All statistical analysis will be conducted using SAS version 9.3 or later (SAS Institute Inc., Cary, NC, USA) or other widely accepted statistical or graphical software as required. All efficacy and safety data endpoints will be summarized using the full analysis set (FAS) defined below as all participants who enter the HP period and use the HP period for any length of time.

Continuous data will be summarized with mean, standard deviation, median, minimum, maximum, and number of evaluable observations. Categorical variables will be summarized with frequency counts and percentages. Confidence intervals may be presented, where appropriate, using the t-distribution for continuous data and Clopper-Pearson Exact method for categorical variables.

Descriptive statistics will be used to summarize data by time point and overall. All safety analyses will be presented using descriptive statistics for the full analysis set. No inferential statistics will be reported.

### Patient and public involvement

No patient was involved in the design of the study.

## Data Availability

All data produced in the present work are contained in the manuscript

## ETHICS AND DISSEMINATION

We hypothesize that the HP treatment in patients with solid malignancies not responding to anti-PD-1 agents will be safe and lead to decreases in the concentrations of EVs and improvements in T cell numbers and activity. If successful, the results of this study will inform the design of a subsequent Pre-Market Approval (PMA) study of the efficacy and safety device in this population.

This trial has been approved by the Human Research Ethics Committees (HREC) of the investigative sites involved. Data is being maintained on a secure database with deidentified participant information. Upon completion of the study, data will be shared in a peer-reviewed journal.

All changes to the protocol which may impact the content of the study and potential benefit or risks for the participants, or which may affect participant safety, including changes of study objectives, study design, patient population or study procedures will be placed in a formal amendment to the protocol and participant information sheet and informed consent forms, and require review and approval by the Ethics Committees at the participating investigative sites prior to implementation.

